# Mechanisms and Pathways Linking Depression and Type 2 Diabetes Outcomes: A Scoping Review

**DOI:** 10.1101/2025.02.21.25322657

**Authors:** Andualem Derese, Sisay Sirgu, Yohannes Gebreegzhiabhere, Charlotte Hanlon

## Abstract

**Aims:** People with diabetes experience a significantly higher prevalence of mental health issues, particularly depression. This adversely affects their diabetes management and overall health. This scoping review aims to develop a conceptual framework for understanding the connection between depression and diabetes outcomes globally, specifically focusing on intermediary factors that may influence this relationship.

**Methods:** PubMed, EMBASE, PsycINFO, and Global Index Medicus were searched using relevant keywords on 17^th^ May 2024. The inclusion criteria encompassed peer-reviewed studies involving adults diagnosed with Type 2 diabetes that assessed depression and analysed its impact on diabetes outcomes through various pathways

**Results:** The review identified 30 studies examining the association between depression and diabetes outcomes. Results indicate that while depression is linked to poorer diabetes outcomes, the mechanisms are complex and often mediated by factors such as self-efficacy, social support, and diabetes-related distress. Notably, self-efficacy emerged as a critical mediator in the relationship between depression and self-management behaviours. Furthermore, social support was identified as a protective factor that can reduce the adverse effects of depression on glycaemic control.

**Conclusions:** Addressing mental health concerns in diabetes care is essential for improving patient outcomes. This review underscores the need for integrated interventions that consider psychosocial factors to enhance self-management and glycaemic control among individuals with Type 2 diabetes. Future research should focus on exploring these relationships in diverse populations to inform tailored strategies for effective diabetes management.

■ Individuals with diabetes experience higher rates of mental health issues, particularly depression, which negatively impacts diabetes management and health outcomes.
■ This scoping review identified 30 studies linking depression to poorer diabetes outcomes and developed a conceptual framework that highlights the complex mechanisms involved, including factors such as self-efficacy, self-management, illness perception and social support.
■ The findings emphasize the importance of addressing mental health in diabetes care

## Introduction

People living with diabetes mellitus (DM) have a considerably higher prevalence of mental health problems, especially depression, compared to those without the disease (1). Factors contributing to this mental health burden include pain and disability arising from complications of the illness, concerns about how the illness will affect them, impacts of DM on relationships and work, as well as the challenges of living with a chronic condition (2). To ensure a person’s overall health and well-being, mental health concerns should be addressed. These include both diagnosed conditions or subclinical symptoms, that may impact their ability to manage their condition effectively (3). These psychosocial conditions include depression, anxiety, disordered eating, and cognitive impairment. Healthcare providers are advised to consider screening for these conditions as part of the initial assessment and at periodic intervals (3).

Depression is a mental health problem that is most often comorbid with diabetes (4–7). In a systematic review of observational studies, 28% of people with type 2 diabetes mellitus (T2DM) suffered from varying degrees of depressive disorders (8). Another systematic review of 20 studies on the epidemiology of diabetes and depression showed a three times higher prevalence of depression among people living with type 1 DM and about a two times higher prevalence among people living with type 2 DM compared to those without diabetes (5). Similarly, another systematic review showed that depression and general distress comorbidity are higher among people living with DM compared to the general population and estimated the risk of developing depression in people with type 2 DM to be increased by 24% (6, 9).

Research has shown that comorbidity between diabetes and depression leads to increased morbidity, functional disability, complications and mortality (4, 10–14). Moreover, the occurrence of diabetes and depression comorbidity significantly increases the cost and burden of diabetes (4, 11). A systematic review of the risk of mortality of people living with DM reported a 1.5 times higher risk of mortality related to depression (12). Furthermore, the paper showed that depression was associated with an increased risk of mortality for all-cause and cardiovascular disease in people living with diabetes (12).

There is a paucity of information regarding the specific mechanisms by which depression negatively impacts the outcomes of individuals with Type 2 DM. However, research suggests that depression can negatively impact self-efficacy, illness perceptions, and overall self-management behaviours (15), which in turn are associated with adverse DM outcomes. Understanding these mechanisms is important for designing effective interventions to address depression and ultimately reduce complications associated with DM.

This scoping review aimed to develop a conceptual framework for understanding the relationship between depression and diabetes outcomes globally. The review considers intermediary factors that may influence the relationship between depression and diabetes outcomes.

## Methods Search strategy

To identify relevant publications, PubMed, EMBASE, PsycINFO, and Global Index Medicus (GIM) databases were searched without restriction on publication date. The search was conducted on 17^th^ May 2024. The search terms comprised key terms and MESH terms for depression, diabetes mellitus, self-efficacy and self-care. Articles were searched using the following terms:

Depression[Title/Abstract] OR “major depressive disorder” [Title/Abstract] OR “Depression” [Mesh] OR “Depressive Disorder” [Mesh] OR “Depressive Disorder, Major” [Mesh] AND “diabetes mellitus, type 2” [Title/Abstract] OR “type 2 diabetes” OR “Diabetes Mellitus, Type 2” [Mesh] OR “Diabetes Mellitus” [Mesh] NOT “Diabetes Mellitus, Type 1” [Mesh] AND “self care” [Title/Abstract] OR “self management” [Title/Abstract] OR “Self-care behaviour” [Title/Abstract] OR “self-care” [Title/Abstract] OR “Self Care” [Mesh] OR “Self-Management” [Mesh] OR “illness cognition” [Title/Abstract] OR “illness cognition” [MeSH Terms] OR “illness perception” [Title/Abstract] OR “illness attribution” [Title/Abstract] OR “explanatory model” [Title/Abstract] OR “illness perception” [MeSH Terms] OR “Self Efficacy” [Title/Abstract] OR “Self-Efficacy” [Title/Abstract] OR “Self concept” [Title/Abstract] OR “Self Efficacy” [Mesh].

### Eligibility criteria

This review included peer-reviewed studies written in English that fulfilled the following criteria.

#### Participants

Adults (age ≥ 18 years) diagnosed with Type 2 DM

#### Exposure

Assessment of depression using either:

- Diagnostic categories of depressive disorders according to established criteria (e.g., DSM-5, ICD-10), or
- Standardised depression scales measure depressive symptoms.

#### Outcomes

Studies assessing the mechanisms or pathways through which depression influences diabetes outcomes. These outcomes include glycaemic control, development of diabetes complications, quality of life, mortality, healthcare utilisation and associated costs, psychosocial outcomes (e.g. social support, anxiety), and functional outcomes.

#### Study design

We included quantitative studies that explicitly analysed the pathways or mechanisms of depression affecting diabetes outcomes. This included analytic observational studies (e.g., cross-sectional, cohort or case-control studies) or randomised controlled trials incorporating a mediation analysis or other methods to explore these pathways.

#### Settings

Studies conducted in any global setting encompassing community and clinical settings.

### Data extraction

The primary author extracted the data using a checklist prepared in advance. Author, publication year, country, setting, study design, sample size, outcomes, measures, and key findings were extracted from each article.

### Data synthesis

Data synthesis was conducted to explore the mechanisms linking depression to key diabetes outcomes. The synthesis specifically focused on the pathways through which depression impacts diabetes outcomes. We extracted and synthesised results from studies employing path analysis and structural equation modelling (SEM) to analyse the relationships among the variables.

Synthesising the included studies’ findings, we developed a preliminary conceptual framework that illustrates the complex interplay between depression, distress, and diabetes outcomes. This framework integrates key concepts and identifies potential causal relationships based on the evidence presented in the literature.

## Results

A total of 2581 articles were identified from the four databases. After removing 544 duplicates, 2037 titles and abstracts were screened, and 117 were selected for full-text review. Of these, 30 articles were eligible and included. Non-eligible articles included brief reports, conference papers, and articles that did not assess mechanisms (Figure 1).

### Results of individual studies

Most studies were from high-income countries (n=21), with half (n=15) from the USA. Nine articles reported on studies conducted in six LMIC countries. Except for four studies (one randomised controlled trial (RCT) and three longitudinal studies), most examined the interplay between depression, self-efficacy and self-care through cross-sectional study designs. Most studies (n=26) were conducted in clinical settings. The publication date ranged from 2004 to 2024. The sample sizes ranged from 99 to 917, and about two-thirds had a sample size of less than 300 participants. Most studies used structural equation modelling (SEM) to assess the mechanisms.

### a. Glycaemic control

Glycaemic control is crucial in managing diabetes and is often assessed using various biomarkers. One of the most common and reliable indicators is haemoglobin A1c (HbA1c) (16). HbA1c is the form of haemoglobin that has bonded with glucose. This test retrospectively assesses average blood sugar control over two to three months (17). The articles reviewed found that self-management (18–20), diabetes distress (21, 22), perceived control (23), self-efficacy (22) and fatalism (22) each had a direct effect on glycaemic control. Diabetes fatalism is a complex psychological cycle characterised by perceptions of despair, hopelessness and powerlessness (24). The relationship between depression and glycaemic control was inconsistent. Except for one study (25) demonstrating a direct link between depression and elevated HbA1c levels, the majority of the included studies did not find significant direct association between depression and glycaemic control (19, 20, 26–29). However, many of these studies reported that depression had an indirect effect on glycaemic control. The mediators of this relationship included self-efficacy (18, 27, 28, 30), self-management (18–20, 25), social support (26), diabetes distress (29) and social comparison (or self-evaluations relative to others in the social environment e.g. peers, family members, media figures)(26).

A study by Azami et. al. found a significant association between depressive symptoms and glycaemic control among men but not among women. This association among men was mediated by diabetes self-efficacy (30).

Diabetes-related distress had a direct effect (21, 22, 29) and an indirect effect on glycaemic control through diabetes self-efficacy, self-care and perceived control (18, 23, 28). Other socioeconomic factors like income and higher social support were also indirectly associated with glycaemic control. Social support had an indirect association with glycaemic control mediated through access to care and processes of care (22). Three factors mediated the association between poverty and glycaemic control: cyclical representation of illness (perception of illness as unpredictable and cyclical), avoidance coping and depressive symptoms (31). One study found that avoidance coping was a full mediator, whereas depressive symptoms and a healthy diet partially mediated the association between education level and glycated haemoglobin (HbA1c) level (31). The association between these socioeconomic factors and glycaemic control was no longer significant when each mediator was considered.

A study by Williams et al. evaluated the relationship between self-determination theory constructs (perceived competence and clinical autonomy support) to glycaemic control, depression and patient satisfaction. The paper reported that autonomy support had an indirect effect on glycaemic control through perceived control (32)

Chiu et al. (33) investigated the influence of social support on the relationship between depression and glycaemic control in older Taiwanese adults. Their findings revealed a significant interaction effect. Among individuals reporting lower family and friend support, depressive symptoms at baseline (T1) positively predicted subsequent HbA1c levels, indicating a potential worsening of glycaemic control with increased depression. However, for participants with strong social support, while depressive symptoms (T1) and HbA1c were correlated at baseline, they did not predict each other three years later. This suggests that social support may buffer the negative consequences of depression on glycaemic control in older adults.

### b. Self-management

Several psychosocial factors, including depression, diabetes distress, self-efficacy, diabetes knowledge, social support and social-ecological support resources, were reported to have direct and indirect associations with either overall self-care score or some of the self-care components (20, 21, 28, 34–39). Depression, in particular, was directly linked to self-care behaviours and indirectly related to self-care behaviours through self-efficacy, diabetes distress, social support by health workers and social support by family and friends (21, 27, 34, 36, 40, 41). However, there were also studies which found no significant association between depression and self-care behaviours (23, 42).

Other intra-individual variables, such as decreased emotional well-being, perceived personal control and perceived situational control, were found to indirectly affect diabetes self-care activities through social-ecological support resources (43). Similarly, Enggarwati et al. discovered a positive bidirectional association between social support and self-care activities. More significant social support led to increased engagement in self-care activities, and in turn, higher levels of self-care activities were associated with stronger social support (38). Al-Amer et al. also showed that in Jordanian adults with type 2 diabetes, social support and depression had a negative correlation, and social support was indirectly related to self-care management behaviour through depression (44). Diabetes fatalism had an indirect association with self-care behaviour through diabetes distress (21, 45).

Hudson et al. reported the effect of emotion and illness cognition on self-care behaviours. They found that participants who had concerns about their diabetes had more depression at six months of follow-up, indicating that cognitions may have a direct effect on emotional representations. Conversely, those participants who had higher baseline depression scores were more likely to believe that their diabetes was unpredictable at six months follow-up, which implies a direct effect from emotions to cognitions. Furthermore, baseline personal control beliefs directly affected adherence to diabetes self-care at six months follow up (42).

A study by Gonzalez et al. examined the relationship between diabetes distress, medication adherence, self-efficacy, and perceived control in individuals with type 2 diabetes. The results indicated that diabetes distress indirectly affected medication adherence, mediated by perceived control and self-efficacy, with only perceived control directly associated with better medication adherence (23). Another study by Jiang et al. found that self-efficacy had a positive indirect effect on self-care behaviour, mediated by depression, with higher self-efficacy leading to lower depression and better self-care behaviour. The strength of this indirect effect might vary depending on age groups (39).

Another study found that self-efficacy mediated the relationship between depression and some components of self-management, like foot care among African Americans, but did not mediate among the Hispanic group (35). Self-efficacy also mediates the relationship between depression and medication adherence (40, 46, 47). Similarly, diabetes distress also has an indirect effect on medication adherence through perceived control and self-efficacy (23). Osborne et al. also found a direct inverse relationship between social support and depression and a direct positive relationship between social support and medication adherence (36). Furthermore, depression also had an indirect relationship with medication adherence through perceived general barriers, perceived side effect barriers, and self-efficacy (48).

On the contrary, Gonzalez et al. reported that diabetes distress had no direct effect on self-care behaviours, suggesting that the impact of diabetes distress on self-care behaviours may be more complex and involve other mediating factors (23). See Table 1 below.

**Table 1.**
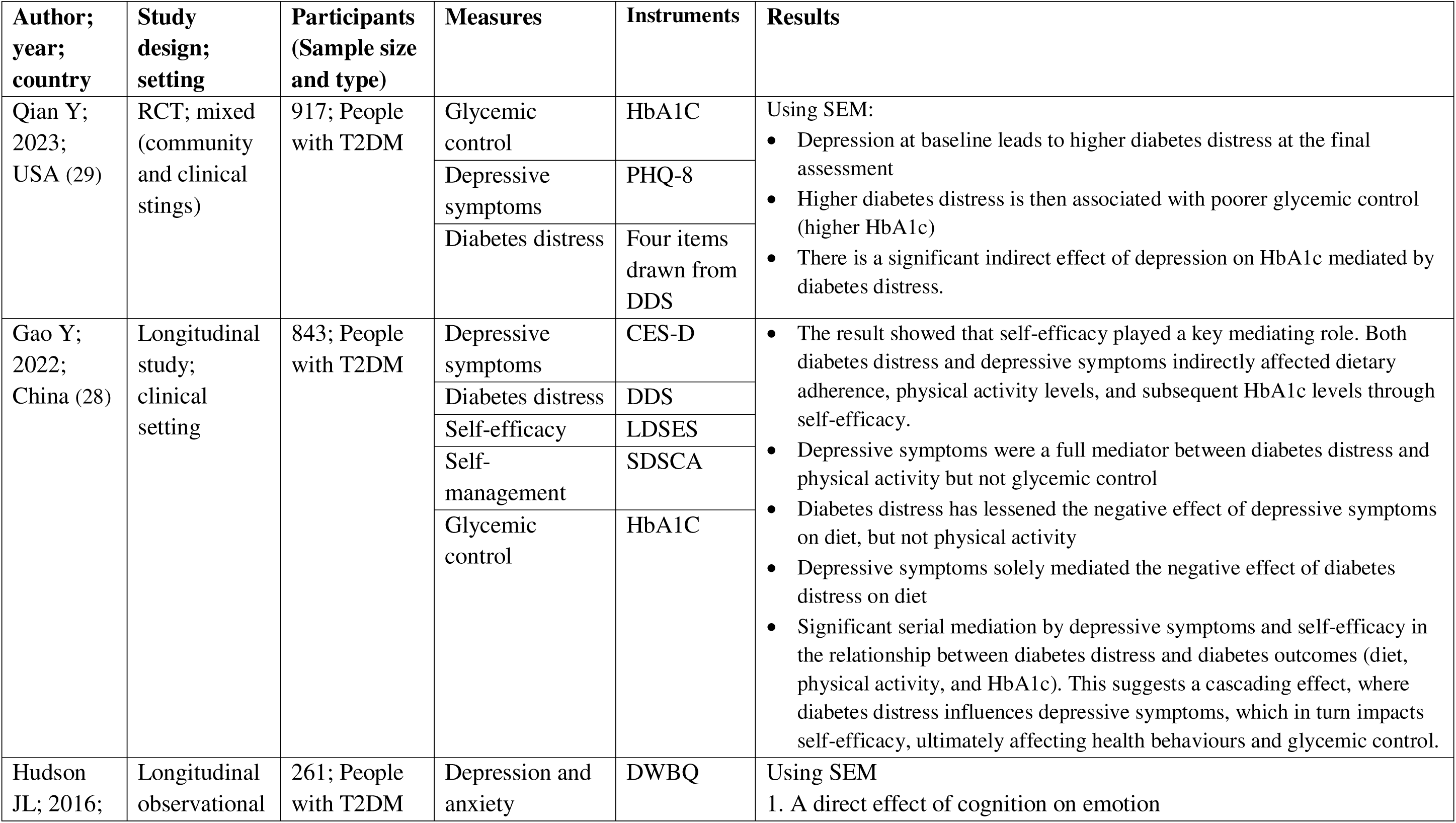
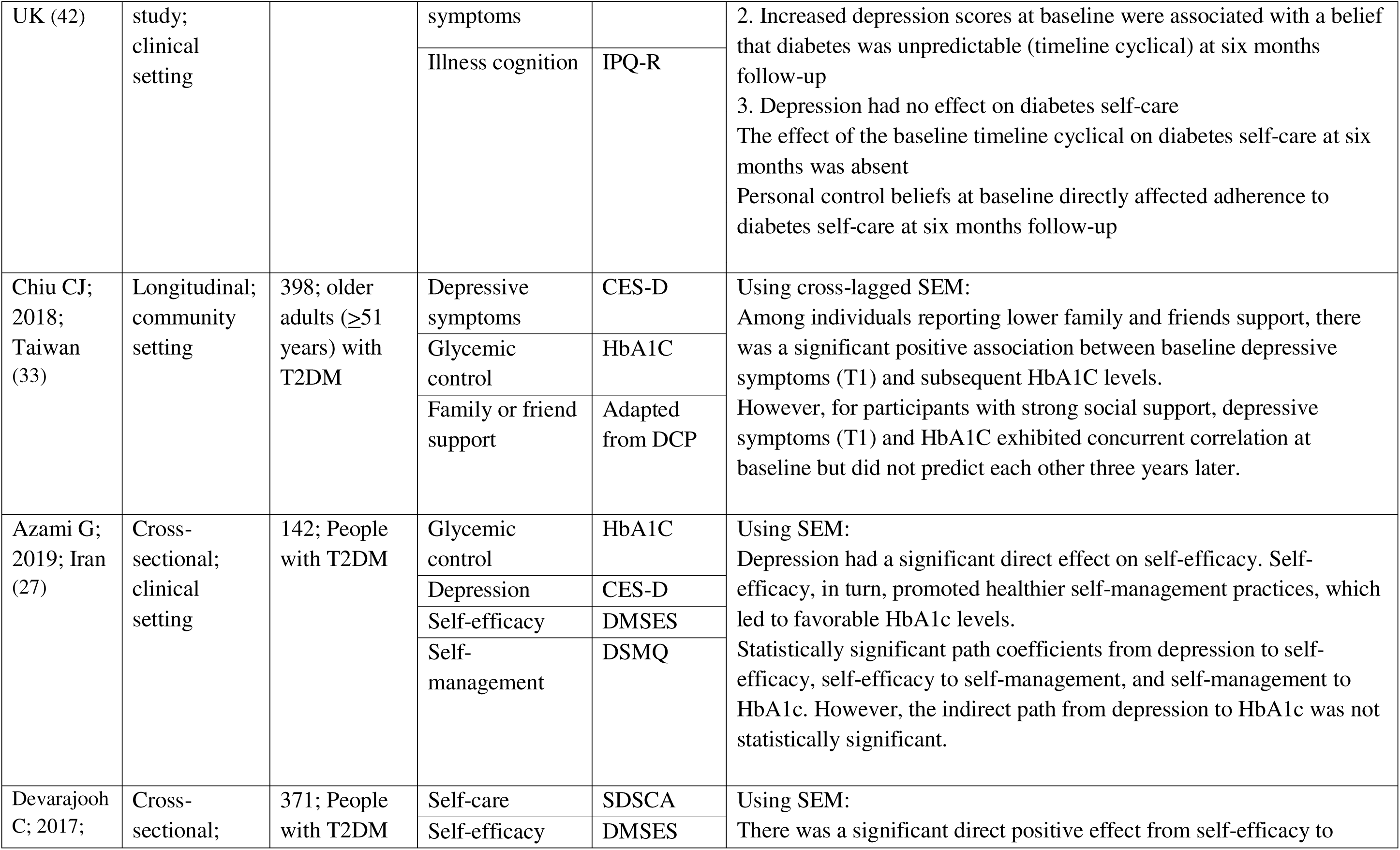
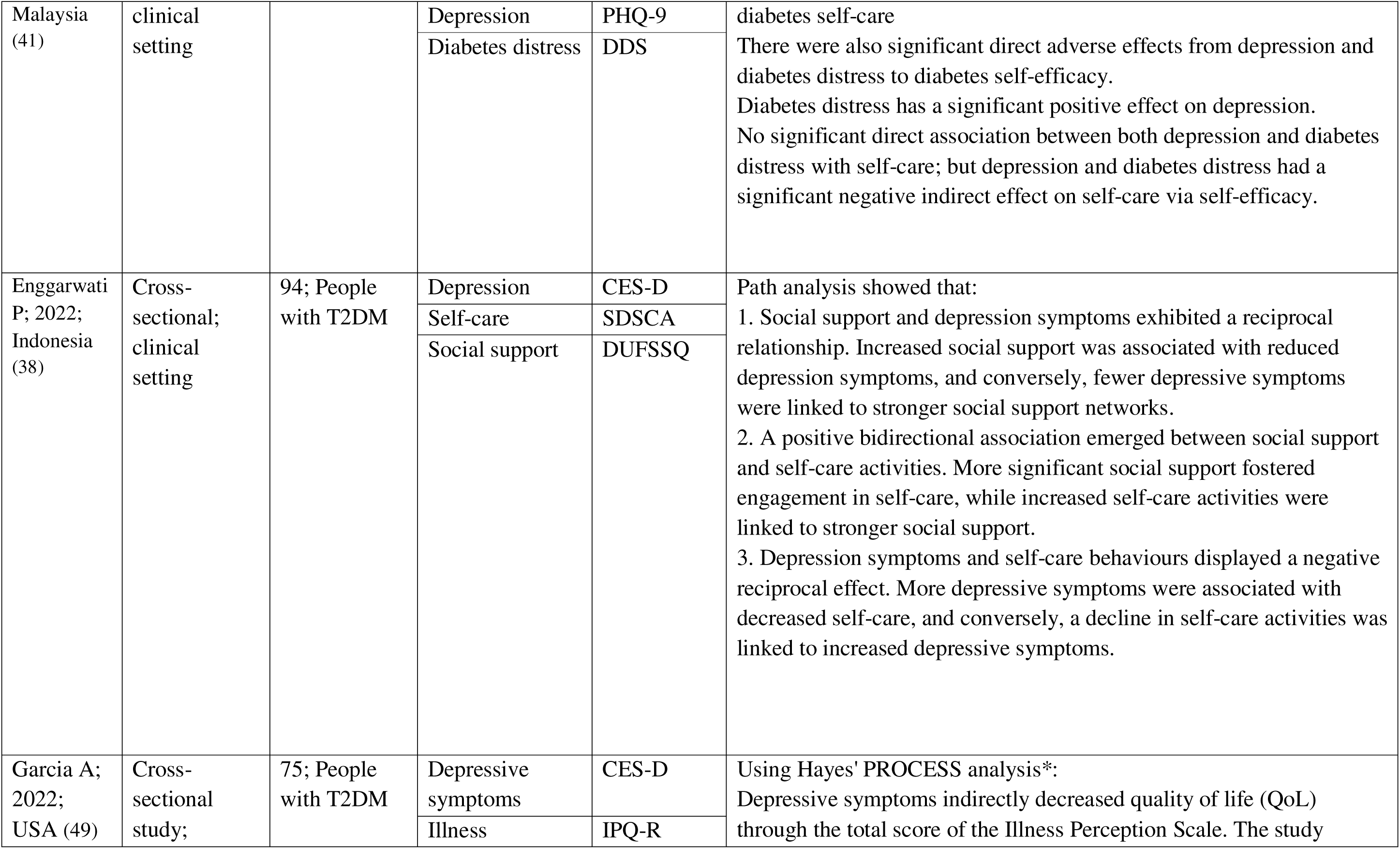
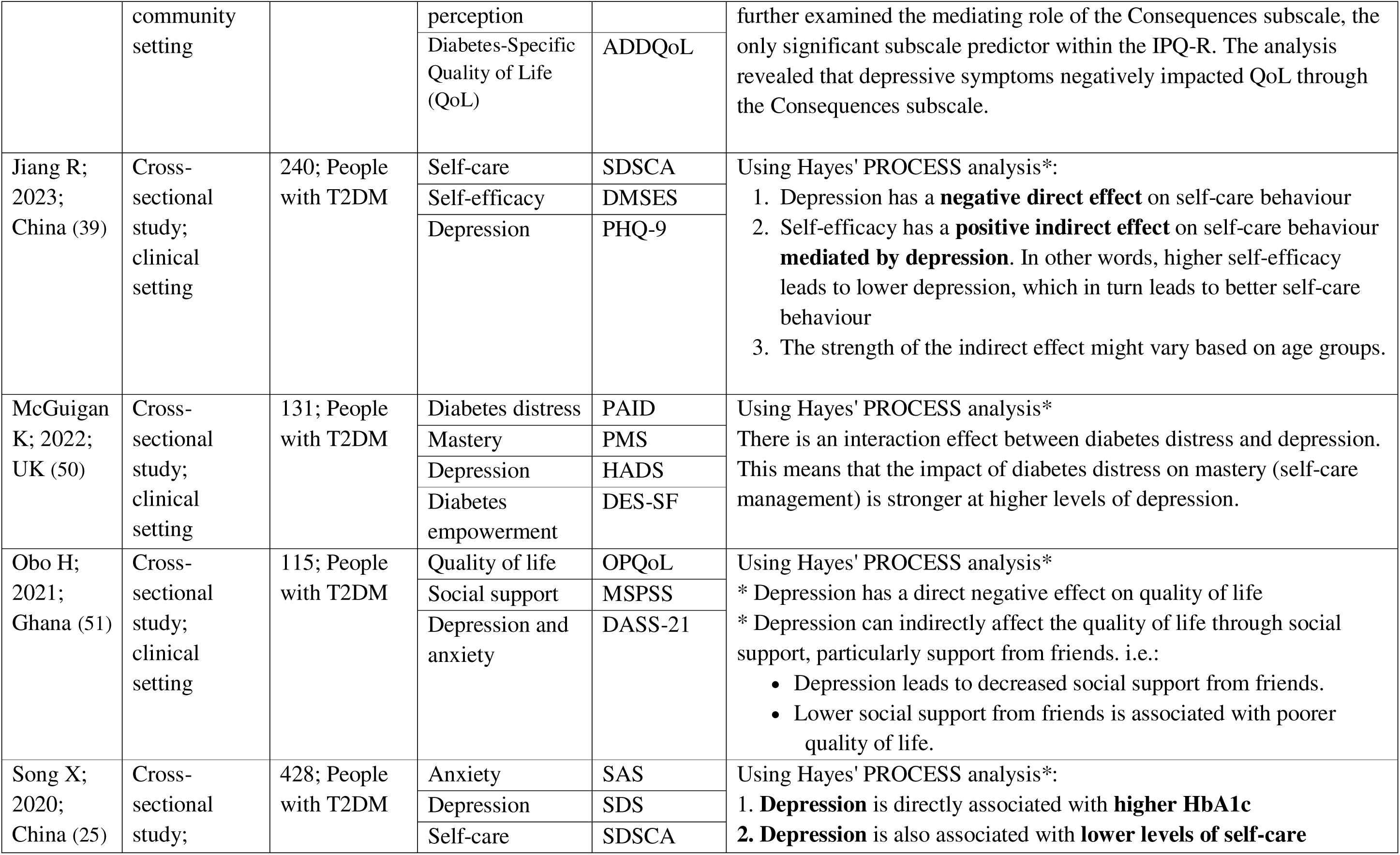
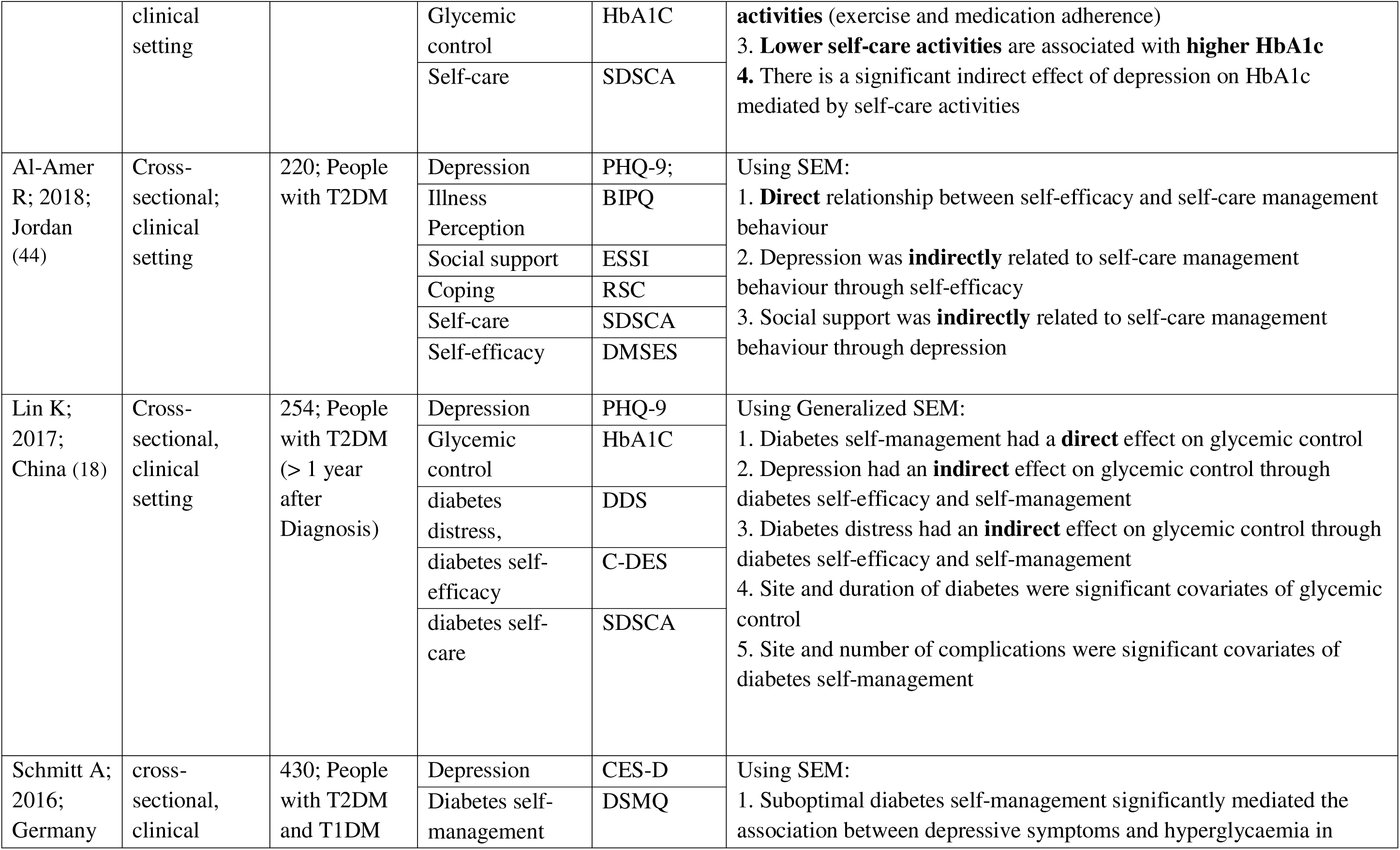
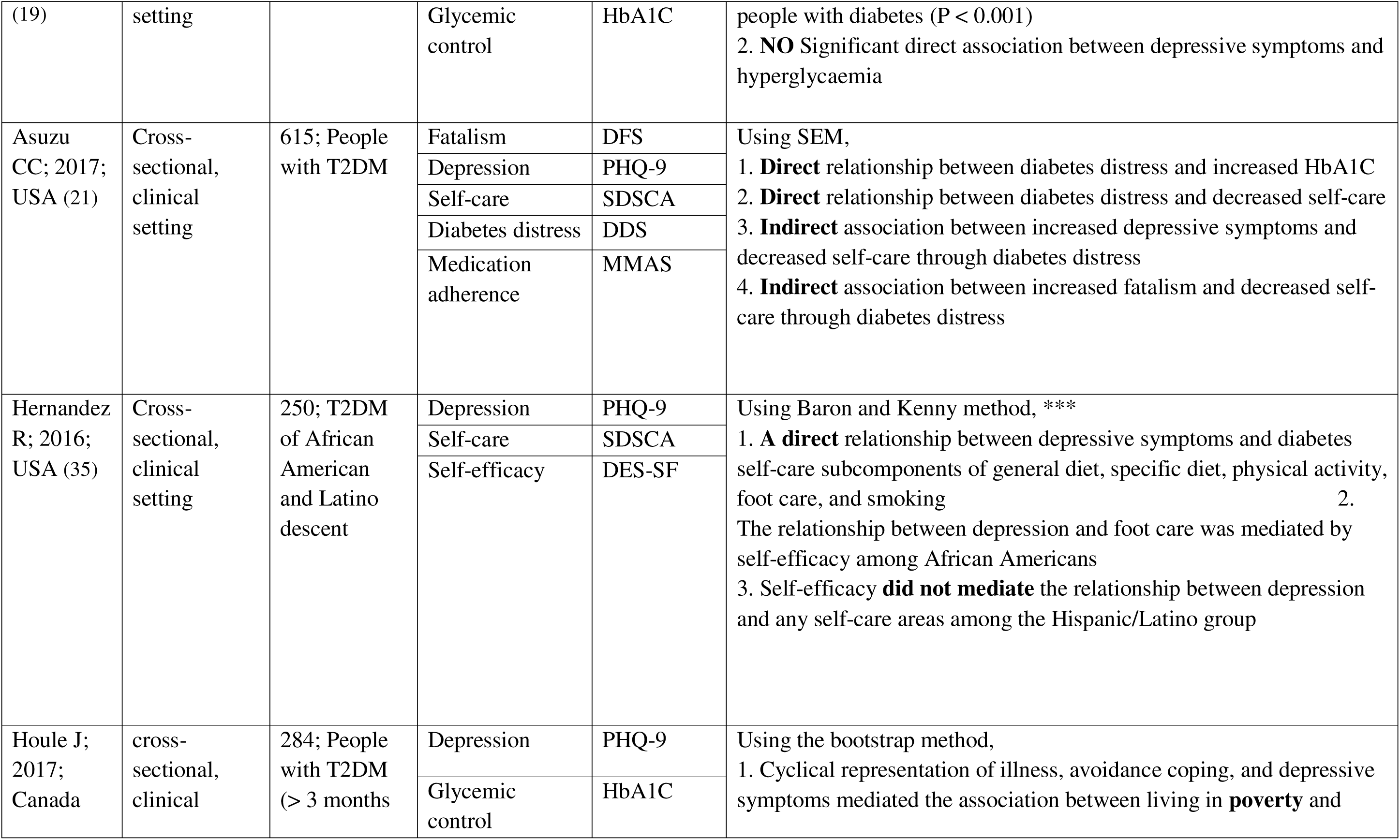
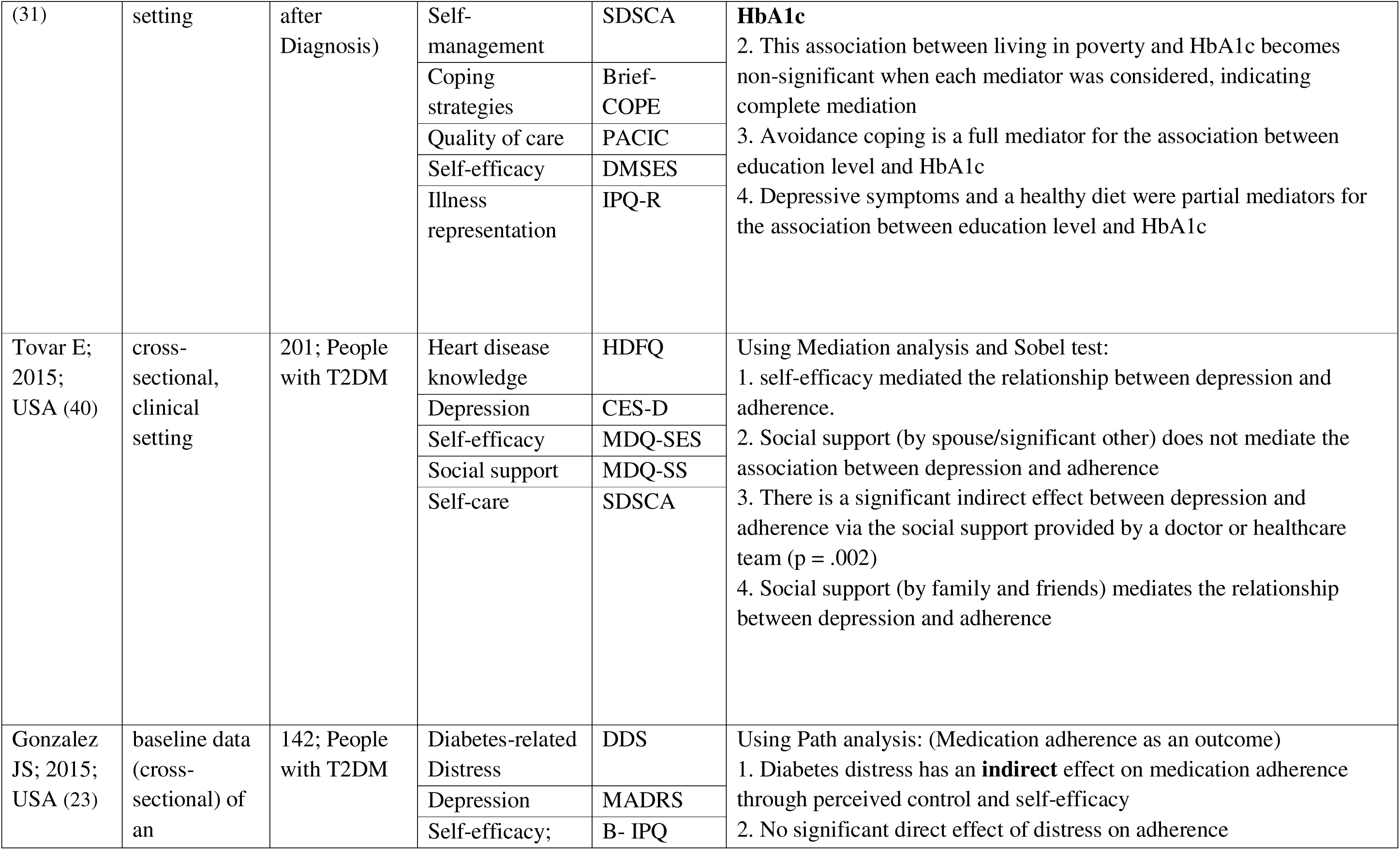
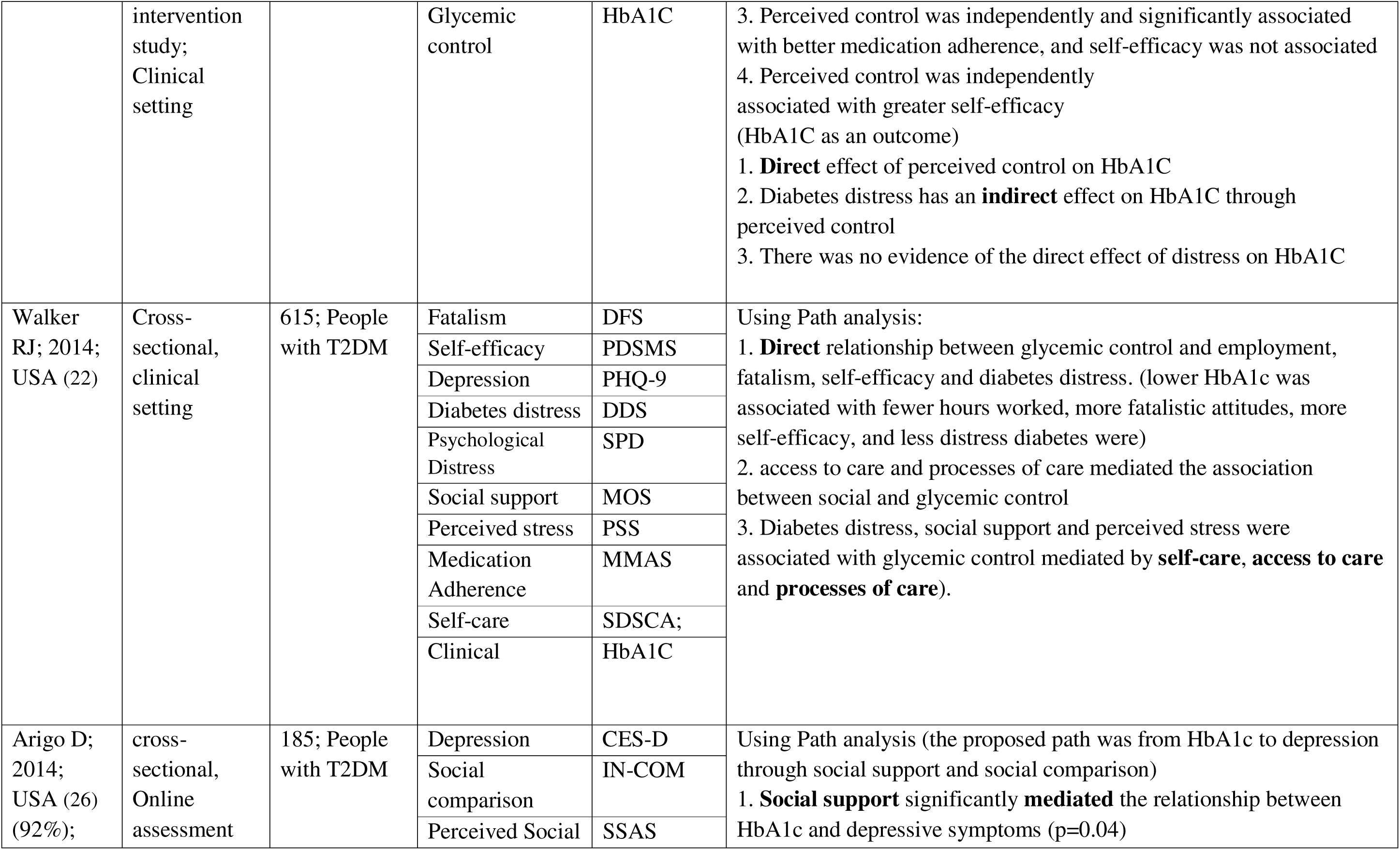
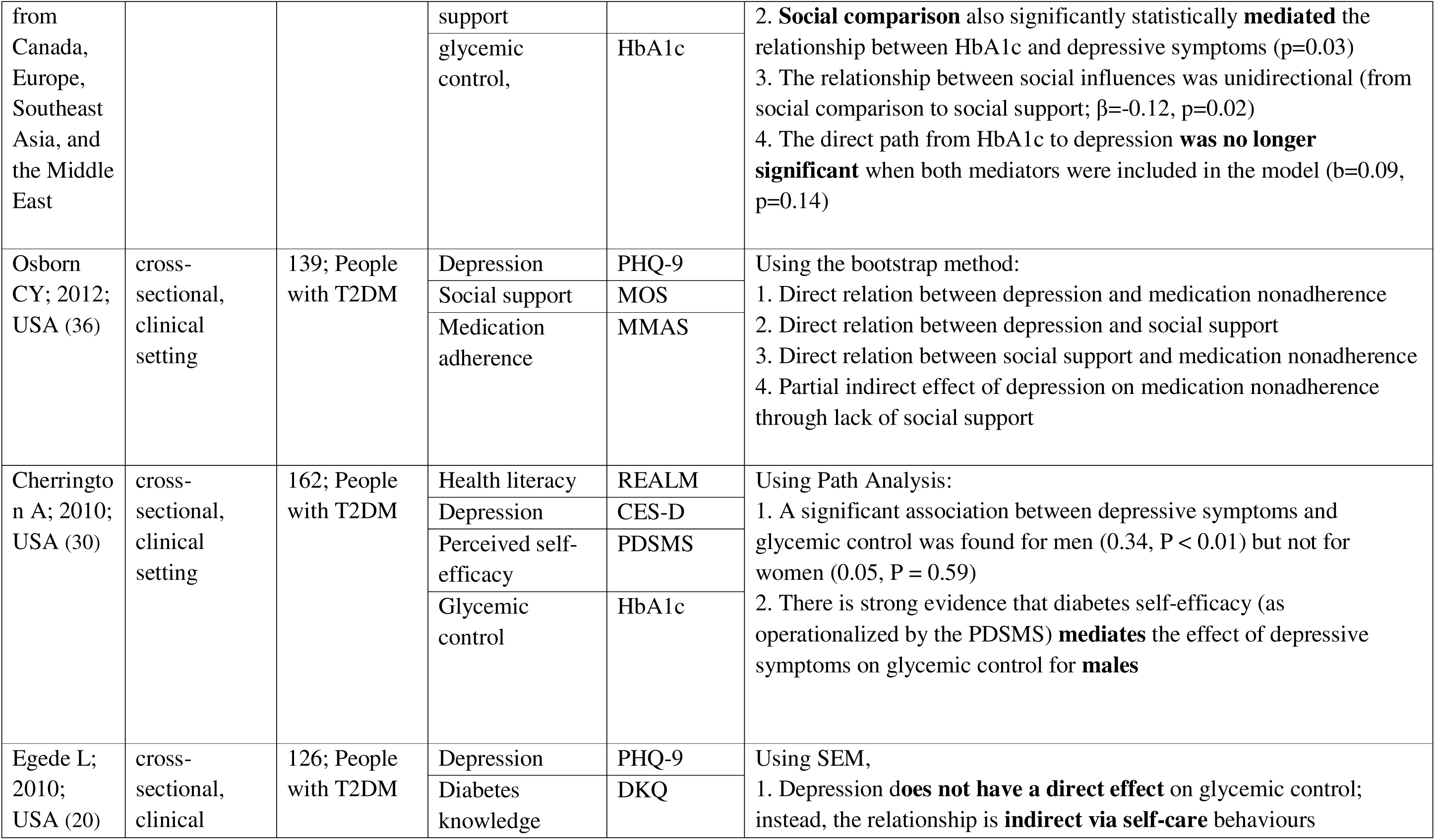
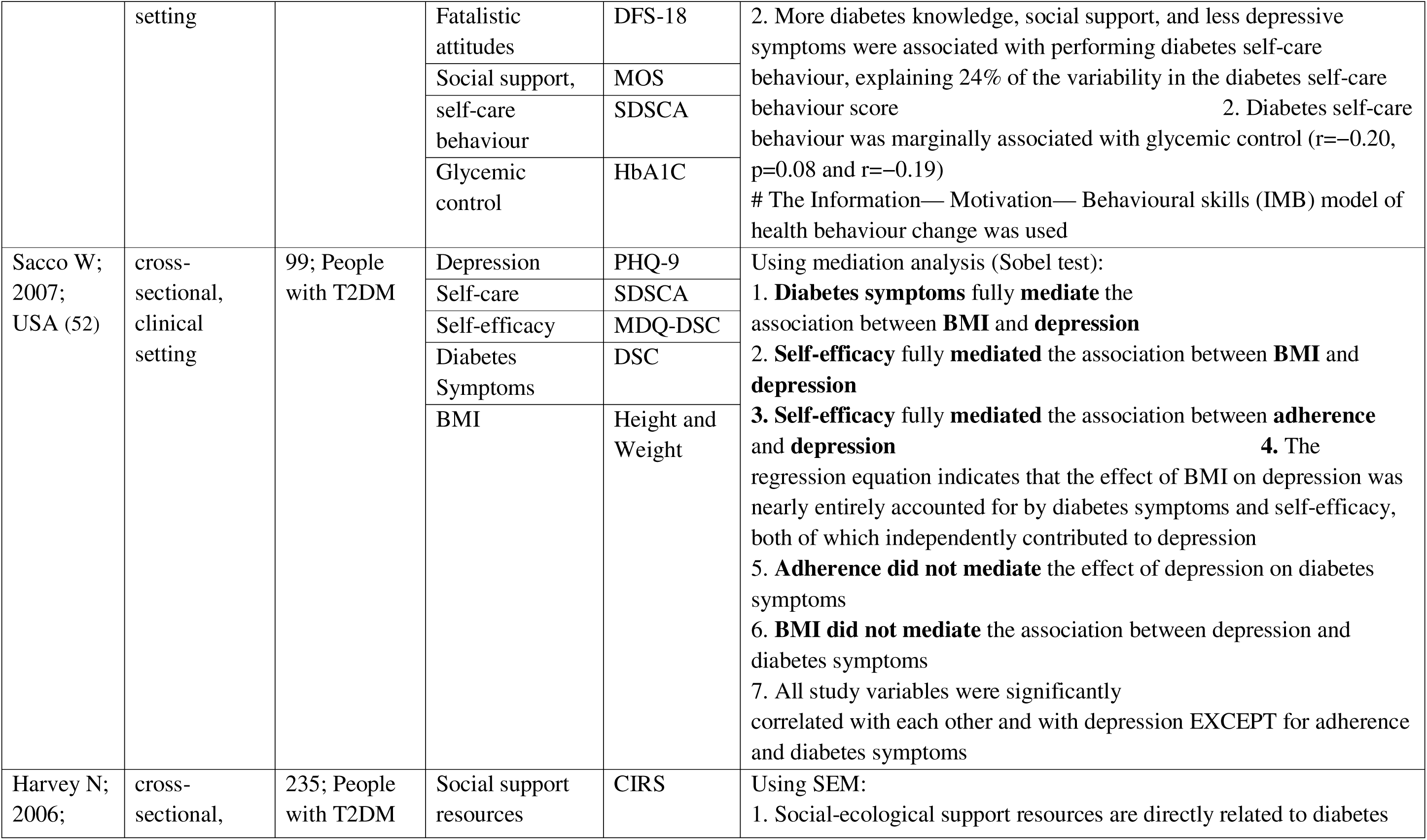
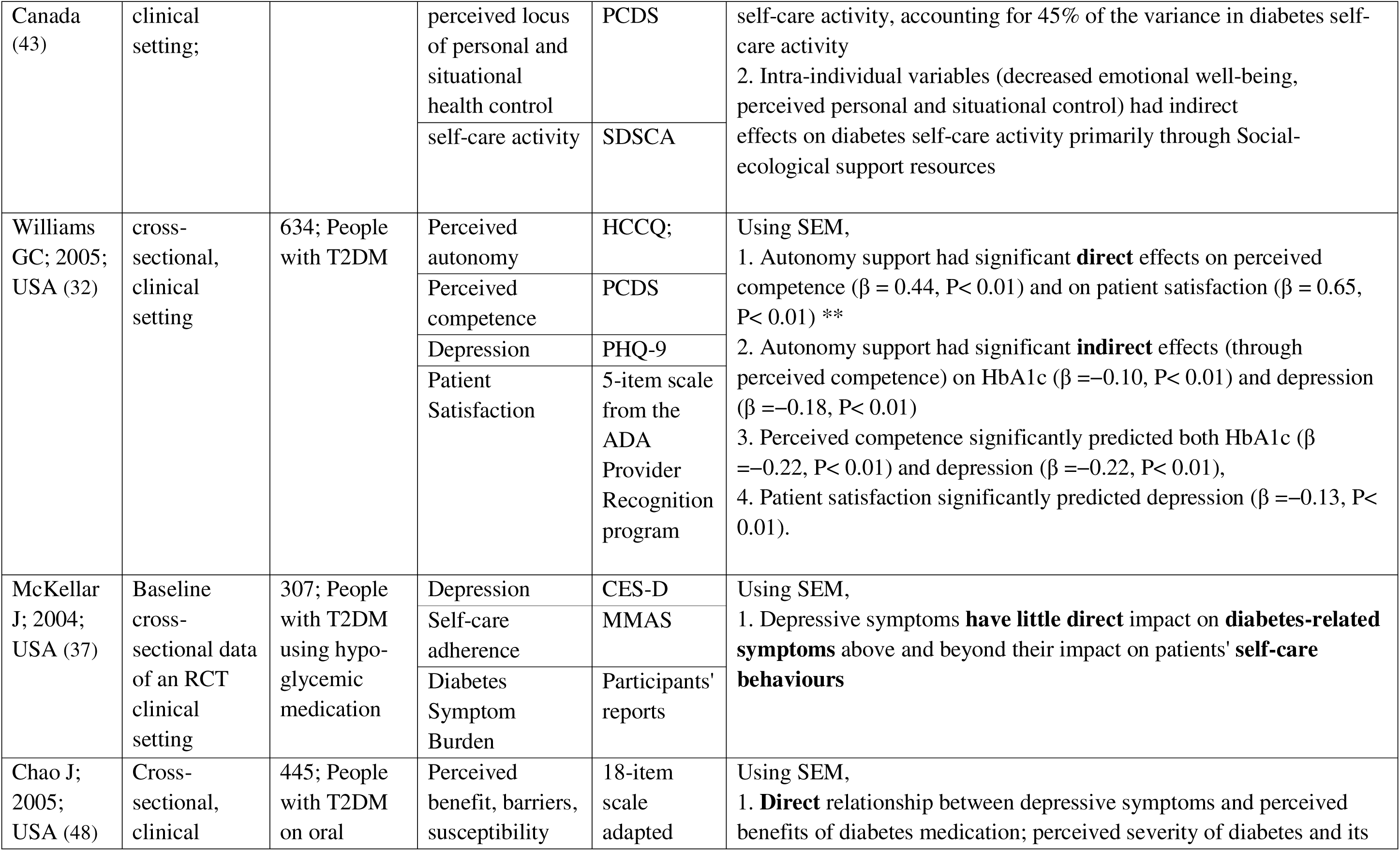
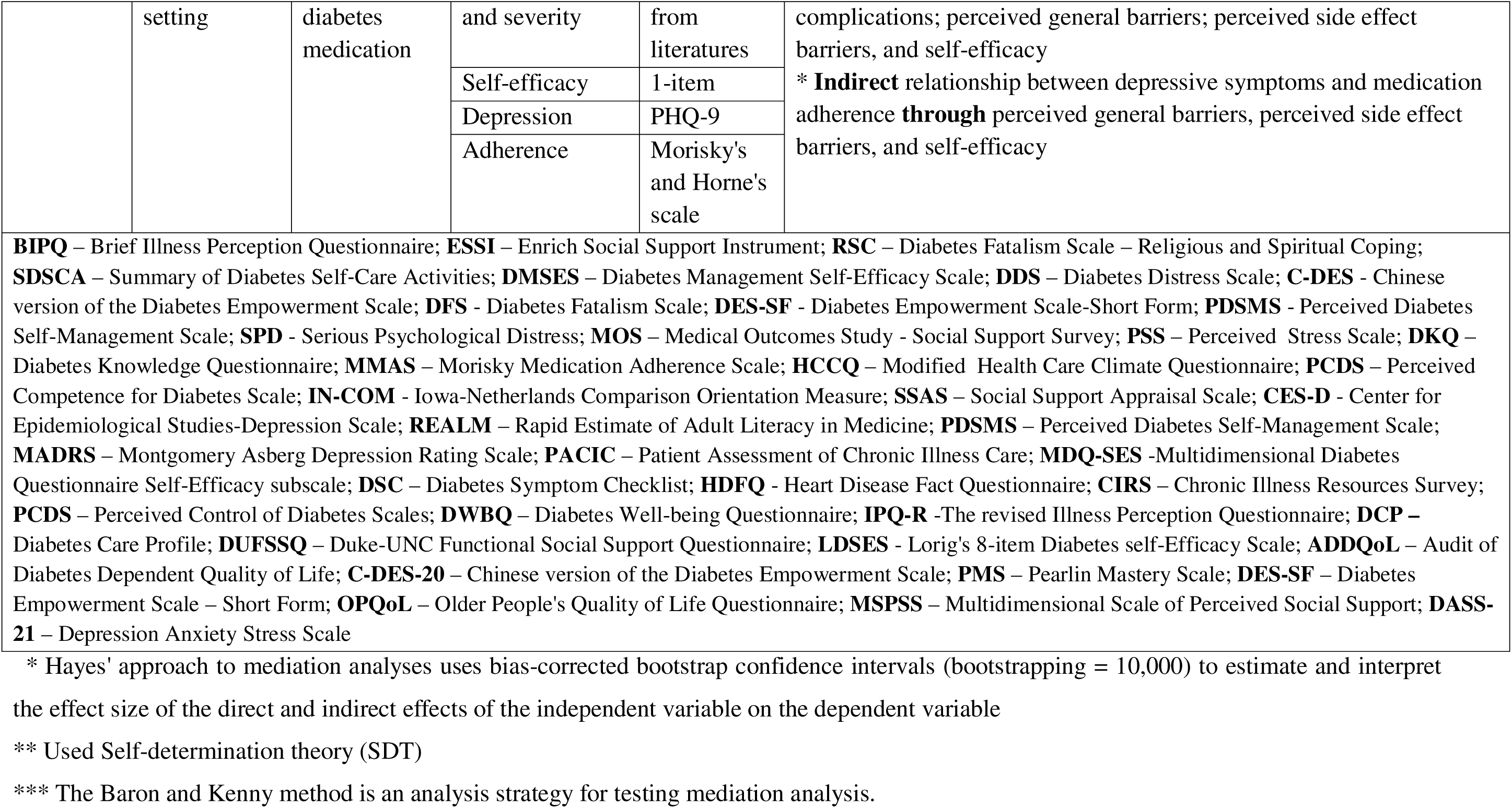
The interrelationships between depression, self-care, self-efficacy and diabetes outcomes.

## Discussion

The results of this scoping review showed the complex relationship between depression and diabetes outcomes, mediated by various psychosocial and behavioural factors, including self-efficacy, illness perceptions, diabetes distress, socioeconomic status and social support. The included studies used various study designs, including longitudinal and cross-sectional approaches across clinical and community settings.

Glycemic control was influenced by depression and diabetes distress across several studies. A randomised controlled trial involving 917 participants with type 2 diabetes mellitus (T2DM) found that depression at baseline led to increased diabetes distress, which was associated with higher HbA1c levels, indicating a significant indirect effect of depression on glycemic control mediated by diabetes distress (29). Similarly, Gao et al. revealed that self-efficacy has important role in mediating the relationship between diabetes distress and glycemic control, suggesting that higher diabetes distress influences depressive symptoms, which subsequently affect health behaviours and glycemic control (28). In contrast, other studies reported that while participants with higher depression scores were more likely to perceive their diabetes as unpredictable, this perception did not directly lead to changes in self-care behaviours or glycemic control (42).

Self-management behaviours were consistently linked to both depression and self-efficacy. Studies demonstrated that depression had a direct negative effect on self-efficacy, which in turn promoted healthier self-management practices (27, 41). Enggarwati et al. emphasised the reciprocal relationship between social support and self-care activities, indicating that increased social support fosters engagement in self-care practices, further strengthening social networks (38). These findings indicated the importance of addressing psychological factors and enhancing social support to improve self-management among individuals with type 2 diabetes.

The studies reviewed demonstrate a clear relationship between depression and diabetes self-efficacy. For example, Azami et al. (27) found a significant direct effect of depression on self-efficacy. Good self-efficacy subsequently promoted healthier self-management practices, leading to favourable HbA1c levels. This is consistent with a study conducted in Malaysia, which indicated that both depression and diabetes distress negatively impacted self-efficacy (41). Hudson et al. reported that participants with higher baseline depression scores were more likely to perceive their diabetes as unpredictable over time (42). This reciprocal relationship aligns with self-efficacy theory, suggesting that emotional states can enhance or undermine self-efficacy (53).

Diabetes distress emerged as a critical mediator in the relationship between depression and glycaemic control. A study by Qian et al. demonstrated that higher levels of diabetes distress mediated the relationship between depression and poorer glycaemic control, indicating a cascading effect where diabetes distress influences depressive symptoms, which in turn affects health behaviours (29). Another study revealed a significant serial mediation by both depressive symptoms and self-efficacy in the relationship between diabetes distress and health outcomes such as dietary adherence and physical activity (28). These findings highlight the necessity of addressing psychological factors such as diabetes distress to improve glycaemic control among individuals with comorbid depression.

Despite these insights, several limitations must be acknowledged. Most included studies employed cross-sectional designs, limiting causal inferences regarding the relationships between variables. Additionally, most studies were conducted in high-income countries, suggesting a need for more research in LMICs to understand the unique challenges faced by these populations. Future research should prioritise longitudinal studies to establish causal relationships and explore community-based interventions that comprehensively address mental health and diabetes management. Additionally, it is important to consider the influence of contextual factors such as socioeconomic status, cultural beliefs, and healthcare access on the relationship between mental health and diabetes self-care. Qualitative studies can further illuminate the mechanisms underlying these relationships and identify culturally appropriate interventions.

In conclusion, this scoping review underscores the complex interplay between depression and diabetes outcomes, mediated by various factors such as self-efficacy, illness perception, and diabetes distress. As a broader contextual factor, socioeconomic status also plays a role in shaping these relationships. Additionally, social support appears to moderate the effects of depression on diabetes outcomes by potentially buffering the negative impact of psychological factors on self-management and quality of life. The findings emphasise the necessity for integrated interventions that address both mental health issues and diabetes management comprehensively. Interventions aimed at enhancing self-efficacy, reducing diabetes distress, and fostering social support networks may be particularly beneficial in improving glycaemic control and overall health outcomes for individuals living with comorbid depression and diabetes.

## Data Availability

All data produced in the present work are contained in the manuscript and supplementary files

