## Supplementary material for "Mechanisms and Pathways Linking Depression and Type 2 Diabetes Outcomes: A Scoping Review": Figure 1_PRISMA flow diagram.docx

**Database search results**

Pubmed: 1000

Embase: 1127

PsycINFO: 352

Global Index Medicus (GIM): 102

Total: 2581

**Titles and abstracts screened**: 2037

**Duplicates removed**: **544**

Excluded based on title and abstract screen: **1940**

**Excluded based on full text screen: 67**

- Protocol (4)
- Conference papers (13)
- Mechanism not assessed (49)
- Full text not found (1)

**Included articles**: 30

Full texts screened for mechanisms: **97**

Figure 1: PRISMA flow diagram for pathways of depression and diabetes outcomes
