## Supplementary material for "Mechanisms and Pathways Linking Depression and Type 2 Diabetes Outcomes: A Scoping Review": Figure 2_Conceptual framework.docx

**Synthesis of findings and development of the conceptual framework**

**Psychosocial factors	Mediating factors	Outcomes**

Income levels

Social support

Self-efficacy

Illness perception

BMI

Diabetes Distress

Medication Adherence

Depression

Glycaemic control

Self-management

Diabetes symptom burden

A

Quality of Life

Figure 2: Conceptual Framework
